# Unraveling the Role of Gut Microbiota and Plasma Metabolites in Fibromyalgia: Insights from Mendelian Randomization and Dietary Interventions

**DOI:** 10.1101/2025.01.02.25319928

**Authors:** Mengqi Niu, Jing Li, Xiaoman Zhuang, Chenkai Yangyang, Yali Chen, Yingqian Zhang, Michael Maes

**Author notes:** Corresponding author: Prof. Dr. Michael Maes, M.D., Ph.D. Sichuan Provincial Center for Mental Health Sichuan Provincial People’s Hospital, School of Medicine, University of Electronic Science and Technology of China Chengdu 610072, China, Michael Maes Google Scholar profile, Prof. Dr. Michael Maes, M.D., Ph.D. https://scholar.google.co.th/citations?user=1wzMZ7UAAAAJ&hl=th&oi=ao Highly cited author: 2003-2023 (ISI, Clarivate), ScholarGPS: Worldwide #1 in molecular neuroscience; #1/4 in pathophysiology, Expert worldwide medical expertise ranking, Expertscape (December 2022), worldwide: #1 in CFS, #1 in oxidative stress, #1 in encephalomyelitis, #1 in nitrosative stress, #1 in nitrosation, #1 in tryptophan, #1 in aromatic amino acids, #1 in stress (physiological), #1 in neuroimmune; #2 in bacterial translocation; #3 in inflammation, #4-5: in depression, fatigue and psychiatry. Mengqi Niu and Jing Li contributed equally to this work.

## Abstract

**Background:** Fibromyalgia (FM) is a complex disorder characterized by chronic pain, fatigue, and functional impairments, with unclear pathological mechanisms. Gut microbiota and plasma metabolites have been implicated in FM, but their causal relationships remain unexplored.

**Objective:** This study aims to assess the causal relationships between gut microbiota, plasma metabolites, and FM using Mendelian randomization (MR) analysis and to explore potential mediating mechanisms.

**Methods:** Public genome-wide association study data were analyzed using bidirectional MR. Associations between gut microbiota, plasma metabolites, and FM were evaluated, and multivariable MR identified mediating metabolites. Results were validated with inverse variance weighted, MR-Egger, and weighted median methods, with metabolic pathway enrichment analysis for further insights.

**Results:** MR identified protective associations between FM and four taxa (family *Enterobacteriaceae*, genus *Butyricicoccus*, genus *Coprococcus1*, and order *Enterobacteriales*) and risk associations with genus *Eggerthella* and genus *Ruminococcaceae UCG005*. Additionally, 82 plasma metabolites linked to pathways such as caffeine metabolism, α-linolenic acid metabolism, GLP-1, and incretin regulation were associated with FM. Mediation analysis revealed *Enterobacteriaceae* and *Enterobacteriales* influenced FM risk through 2,3-dihydroxypyridine and palmitoylcholine.

**Conclusion:** Personalized dietary interventions, such as limiting caffeine intake, increasing omega-3 fatty acid consumption, adopting a low glycemic index diet, and reducing the intake of high-oxalate foods, may effectively alleviate FM-related symptoms by modulating metabolic pathways, reducing inflammation, and mitigating oxidative stress. This study highlights the intricate interactions between the gut microbiota and metabolic pathways, providing critical scientific evidence and actionable targets for clinical interventions, dietary management, and precision medicine approaches in FM treatment.

## 1. Introduction

Fibromyalgia (FM) is a long-term disorder characterized by diffuse pain across the musculoskeletal system, accompanied by stiffness in localized areas and heightened pressure sensitivity, commonly known as tender points within soft tissues ^1^. While pain serves as the defining and most prominent symptom of FM, the condition also involves various other manifestations, such as chronic fatigue, disrupted sleep patterns, and functional impairments—symptoms often without identifiable structural or pathological causes ^2^. FM impacts around 2.7% of the global population, with its prevalence being significantly higher in women, individuals over 50 years of age, people from lower socioeconomic backgrounds, and those classified as obese ^3^. Despite extensive research, the exact pathogenic mechanisms driving FM remain unclear, and effective, universally accepted treatment options are yet to be developed ^2^.

Previous studies, including one conducted in 2007, have proposed a link between FM symptoms observed in chronic fatigue syndrome (CFS) and the increased translocation of Gram-negative gut bacteria, suggesting the presence of a "leaky gut" ^4^. In a similar vein, FM symptoms in individuals with schizophrenia have been associated with paracellular pathway disruptions and increased bacterial translocation ^5^. Such evidence suggests that gut dysbiosis could contribute significantly to the development and progression of FM. For example, studies conducted by Minerbi et al. ^6^ along with further research by Minerbi and Fitzcharles et al. ^7^, have identified notable associations between gut microbiota and FM. Moreover, a systematic review has emphasized the significant role of intestinal microbiota in the pathophysiology of FM ^8^.

FM is a chronic disorder primarily characterized by abnormal pain processing in the central nervous system. Recent studies, however, suggest that systemic metabolic alterations may play a critical role in its pathophysiology. Immune and metabolomics research has revealed significant abnormalities in multiple pathways in FM patients, including those related to the immune system, metabolism, oxidative stress, muscle energy and general energy metabolism, and mitochondrial functions ^9–13^. Numerous metabolic problems have been identified in FM concerning amino acid metabolism, including tryptophan, branched-chain amino acids, glutamate, and gamma-aminobutyric acid, lipid metabolism, and protein metabolism, including decreased levels of peptidases ^12–14^. Additionally, imbalances in polyunsaturated fatty acids (PUFAs) contribute to the development of chronic pain syndromes ^9^. For example, in chronic fatigue syndrome, the omega-3/omega-6 ratio was inversely associated with fibromyalgia symptoms ^15^. Furthermore, studies have shown that mitochondrial reactive oxygen species (ROS) levels are elevated in the blood mononuclear cells of FM patients and are positively correlated with inflammatory markers, such as tumor necrosis factor-alpha ^10^. Overall, the above results indicate that muscle energy depletion and metabolic disorders, mitochondrial dysfunction, oxidative stress, and immune disbalances may be significantly associated in FM patients.

Although these findings are compelling, the causal relationship among gut microbiota, plasma metabolites, and FM remains to be fully elucidated. In recent years, Mendelian randomization (MR) has gained recognition as a robust analytical tool for determining the causal effects of specific exposures on targeted outcomes ^16^. MR utilizes genetic variations, including single nucleotide polymorphisms (SNPs), as instrumental variables to investigate target exposures ^17^. By relying on the random assortment of genetic variations during meiosis, this method effectively minimizes the influence of confounding factors and the risk of reverse causation. The integration of summary statistics from genome-wide association studies (GWAS) within two-sample MR designs significantly strengthens the statistical power for deriving causal inferences ^18^.

Consequently, the present study utilizes GWAS-derived summary data to perform MR analyses, aiming to explore a hypothesized causal link between gut microbiota and FM, mediated by plasma metabolites.

### 2. Materials and Methods

#### 2.1 Study Design

The data used in this analysis were obtained from publicly accessible shared repositories, thus no additional ethical approval was required for this study. First, we examined the causal relationship between the gut microbiome and FM using bidirectional MR, selecting relevant *gut microbiota*. Next, we used the same approach to explore the causal relationship between 1,400 plasma metabolites and FM, identifying significant metabolites. We then applied the same methodology to examine the causal relationship between significant *gut microbiota* and selected metabolites. Finally, mediation MR (multivariable MR) was conducted to further investigate the role of specific plasma metabolites in mediating the relationship between *gut microbiota* and FM (see **Figure 1**).

**Figure 1.**
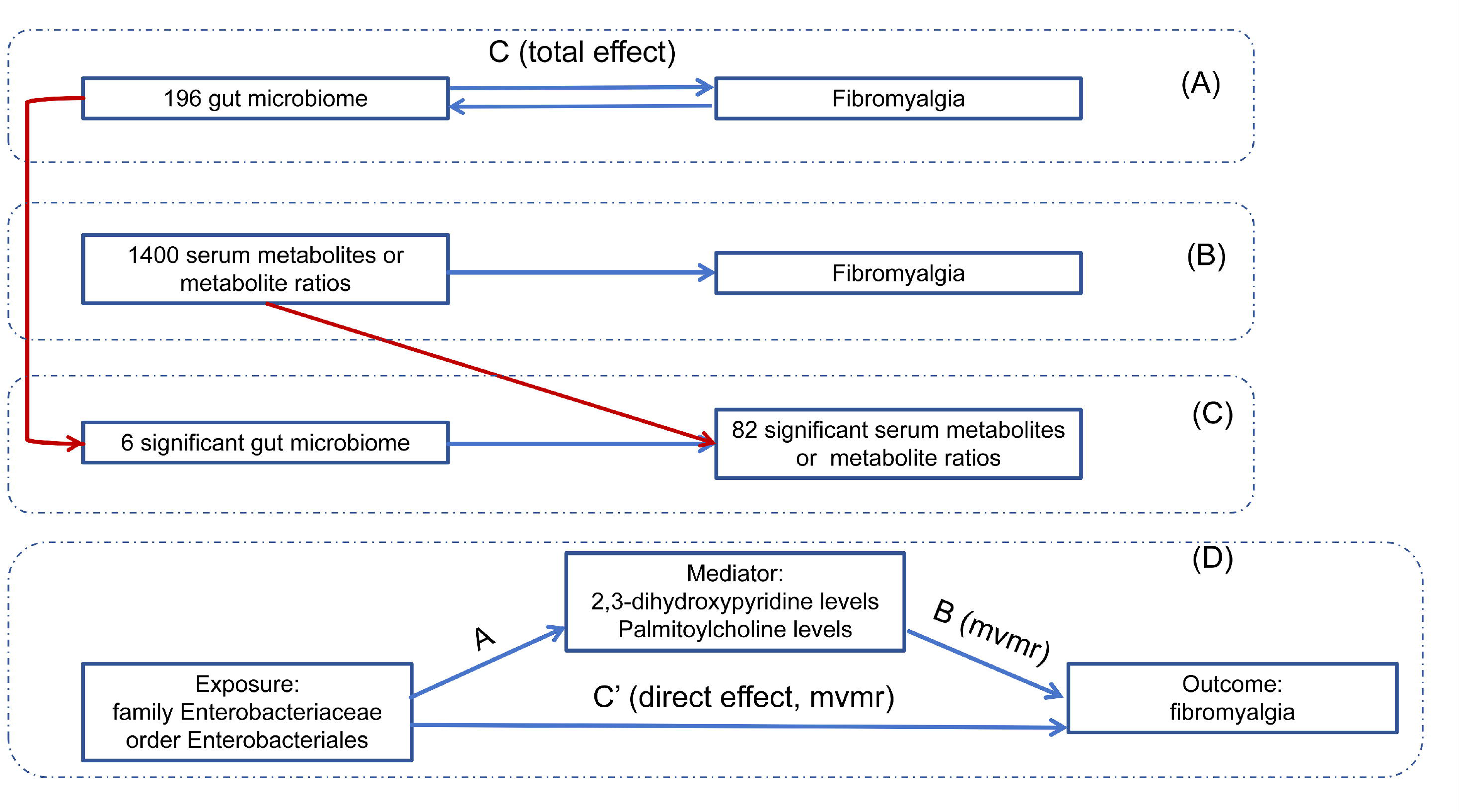
Overall design ideas for this study. (A) Bidirectional mendelian randomization (MR) analysis between Gut Microbiota (GM) and Fibromyalgia (FM). C refers to the total effect using genetically predicted GM as the exposure and FM as the outcome. (B) MR analysis of serum metabolites or metabolite ratios as exposure and FM as outcome. (C) MR analysis of GM (significant GM from Figure 1A) as exposure and serum metabolites or metabolite ratios (significant results from Figure 1B) as outcome. (D) Mediation analysis of GM on Fibromyalgia (FM) through metabolites; C’ represents the direct effect of GM on FM after applying the MVMR method; A is the causal effect of GM on metabolites; B is the causal effect of metabolites on FM after applying the MVMR method; Indirect effect = A * B; Mediation effect = (A * B) / C.

#### 2.2 Data Sources

This study used only aggregated data from European populations to reduce biases due to population heterogeneity. SNPs associated with the human gut microbiome composition were obtained from the latest data by the MiBioGen consortium (https://mibiogen.gcc.rug.nl/). This GWAS included 211 microbial taxa, categorized at six taxonomic levels, from phylum to species ^19^. We obtained data on 1,400 plasma metabolites from Chen et al., including 1,091 blood metabolites and 309 metabolite ratios ^20^. FM-related outcome data were collected from the FinnGen consortium (2023 release, https://r10.finngen.fi/).

#### 2.3 Genetic Instrument Selection

1. We applied a significance threshold of p<1×10^−5^ to ensure a moderate number of SNPs for all *gut microbiota* and plasma metabolites.
2. To confirm the independence of the exposure instruments, we excluded SNPs in linkage disequilibrium (LD; r^2^<0.001, clumping window = 10,000 kb) ^21^.
3. Palindromic SNPs with moderate minor allele frequencies were removed ^22^.
4. To assess potential bias from weak instrumental variables, we calculated F-statistics; an F-statistic above 10 indicated minimal risk of weak instrument bias ^23^.

#### 2.4 MR Analysis

Three MR methods—MR-Egger, weighted median, and inverse variance weighted (IVW)—were applied to analyze causal relationships among the *gut microbiome*, plasma metabolites, and FM. Heterogeneity and pleiotropy were assessed using Cochran’s Q test, MR-Egger intercept test, and MR-PRESSO (Mendelian Randomization Pleiotropy RESidual Sum and Outlier) analysis. A Cochran’s Q test p-value < 0.05 indicated heterogeneity in the results ^24^. The MR-Egger intercept was used to detect directional pleiotropy and bias from invalid instruments ^25^. Finally, MR-PRESSO was conducted to identify outliers that may exhibit horizontal pleiotropy ^26^.

Causal relationships among gut microbiota, plasma metabolites, and FM were determined by:

1. A significant IVW p-value (p < 0.05);
2. Consistency in direction and magnitude across the three MR methods;
3. Passing sensitivity analysis.

In addition, we conducted enrichment analysis of the identified metabolites using the Kyoto Encyclopedia of Genes and Genomes (KEGG), Reactome, Wikipathways, and Resource for Metabolite Pathway Database (RaMP-DB) through the MetaboAnalyst 6.0 platform (https://www.metaboanalyst.ca/) and Wikipathways (https://www.wikipathways.org/).

#### 2.5 Mediation Analysis

Mediation analysis was conducted using multivariable MR to decompose the total effects into direct and indirect (mediated) effects ^27^. The total impact of the gut microbiome on FM was divided into direct effects and indirect effects mediated by plasma metabolites. The mediation proportion was calculated by dividing indirect effects by total effects.

Statistical analyses were performed in R (version 4.3.2) using the MendelianRandomization, MR-PRESSO, and TwoSampleMR packages. For binary outcomes, odds ratios (OR) and 95% confidence intervals (CI) quantified causal effects. For continuous outcomes, results were reported as beta coefficients (β), standard errors (SE), and 95% CI.

#### 3. Results

##### 3.1 Gut Microbiome and Fibromyalgia Causal Relationship

As shown in **Figure 2A**, six gut microbiota taxa showed a causal effect on FM through the three MR methods, without evidence of heterogeneity or pleiotropy. As shown in **Figure 2B**, higher levels of family *Enterobacteriaceae*, genus *Butyricicoccus,* genus *Coprococcus1*, and order *Enterobacteriales* were associated with a reduced risk of FM, suggesting potential protective roles. Conversely, higher levels of genus *Eggerthella* and genus *Ruminococcaceae* UCG005 increased FM susceptibility, indicating possible risk factors. No reverse causality was detected among these associations (**Table S1**).

**Figure 2.**
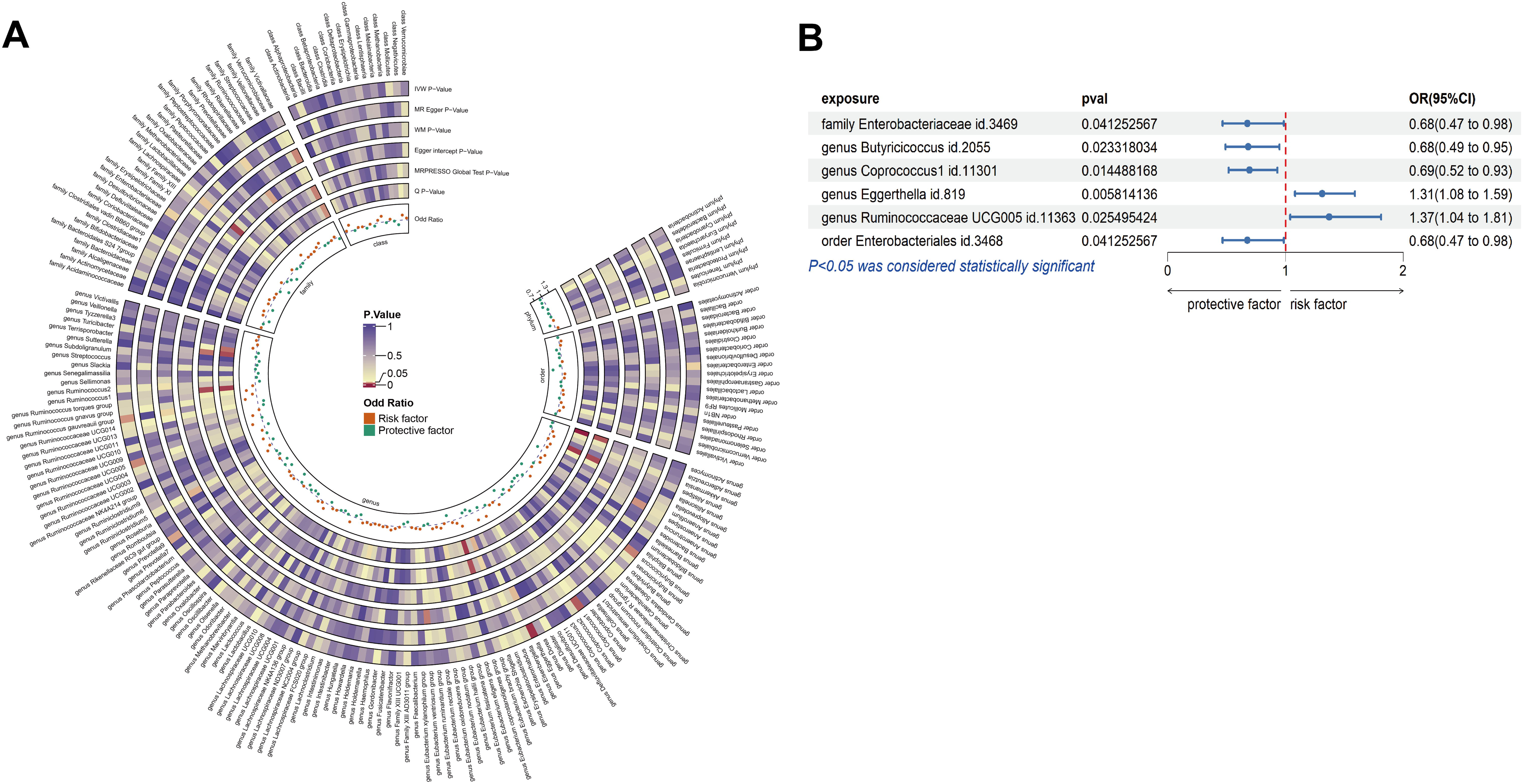
(A) Circular heatmap of mendelian randomization analysis on the causal relationship between gut microbiota and fibromyalgia. (B) Forest plot of causal relationships between significant gut microbiota and fibromyalgia.

##### 3.2 MR Results for Plasma Metabolites and Fibromyalgia

As shown in **Figure 3A**, a total of 82 plasma metabolites associated with FM were identified using three MR analysis methods. These metabolites were selected based on their significance, with no evidence of heterogeneity or pleiotropy. Among the 82 metabolites, 44 were classified as known metabolites, 24 were metabolite ratios, and 14 were unknown metabolites. **Figure 3B** illustrates the causal relationships between the 44 known metabolites and FM, which include 10 amino acids, 2 carbohydrates, 2 cofactors and vitamins, 19 lipids, 2 nucleotides, and 9 exogenous substances. The causal associations of the 24 metabolite ratios and 14 unknown metabolites with FM are detailed in Table 1.

**Figure 3.**
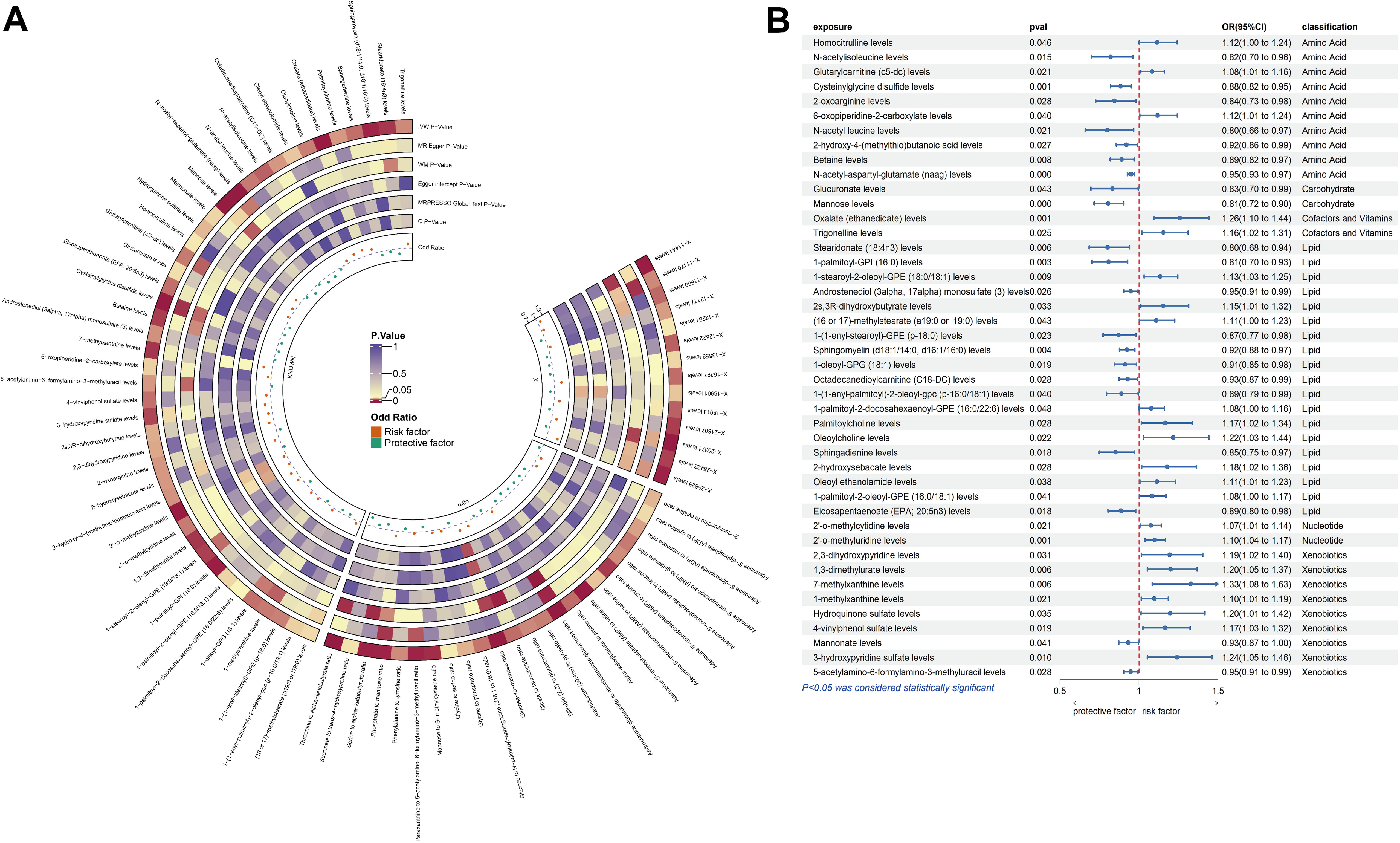
(A) Circular heatmap of mendelian randomization analysis on the causal relationship between significant plasma metabolites and fibromyalgia. (B) Forest plot of causal relationships between significant known metabolites and fibromyalgia.

**Table 1.** Causal effects of plasma metabolite ratios and unknown metabolites on fibromyalgia.

| **Exposure** | **P** | **OR** | **Lower 95% CI** | **Upper 95% CI** |
| --- | --- | --- | --- | --- |
| 2'-deoxyuridine to cytidine ratio | 0.035 | 0.894 | 0.805 | 0.992 |
| Adenosine 5'-diphosphate (ADP) to cytidine ratio | 0.042 | 1.140 | 1.004 | 1.293 |
| Adenosine 5'-diphosphate (ADP) to mannose ratio | 0.029 | 1.139 | 1.013 | 1.282 |
| Adenosine 5'-monophosphate (AMP) to glutamate ratio | 0.021 | 0.809 | 0.676 | 0.969 |
| Adenosine 5'-monophosphate (AMP) to leucine ratio | 0.025 | 0.809 | 0.672 | 0.974 |
| Adenosine 5'-monophosphate (AMP) to proline ratio | 0.007 | 0.822 | 0.712 | 0.948 |
| Adenosine 5'-monophosphate (AMP) to serine ratio | 0.037 | 0.824 | 0.687 | 0.989 |
| Adenosine 5'-monophosphate (AMP) to valine ratio | 0.001 | 0.770 | 0.661 | 0.897 |
| Androstenediol (3α, 17α) monosulfate (3) levels | 0.026 | 0.863 | 0.764 | 0.975 |
| Androsterone glucuronide to etiocholanolone glucuronide ratio | 0.000 | 1.132 | 1.068 | 1.201 |
| Arachidonate (20:4n6) to pyruvate ratio | 0.022 | 0.869 | 0.771 | 0.980 |
| Bilirubin (Z,Z) to glucuronate ratio | 0.024 | 0.914 | 0.845 | 0.988 |
| Citrate to taurocholate ratio | 0.018 | 1.192 | 1.031 | 1.379 |
| Glucose-to-mannose ratio | 0.000 | 0.839 | 0.724 | 0.973 |
| Glucose to N-palmitoyl-sphingosine (d18:1 to 16:0) ratio | 0.020 | 1.238 | 1.100 | 1.394 |
| Glycine to phosphate ratio | 0.034 | 1.075 | 1.006 | 1.149 |
| Glycine to serine ratio | 0.046 | 1.078 | 1.001 | 1.161 |
| Mannose to S-methylcysteine ratio | 0.005 | 0.817 | 0.710 | 0.940 |
| Paraxanthine to 5-acetylamino-6-formylamino-3-methyluracil ratio | 0.004 | 1.052 | 1.017 | 1.088 |
| Phenylalanine to tyrosine ratio | 0.025 | 0.877 | 0.782 | 0.984 |
| Phosphate to mannose ratio | 0.004 | 1.185 | 1.057 | 1.329 |
| Serine to alpha-ketobutyrate ratio | 0.003 | 0.782 | 0.664 | 0.921 |
| Succinate to trans-4-hydroxyproline ratio | 0.045 | 0.874 | 0.767 | 0.997 |
| Threonine to alpha-ketobutyrate ratio | 0.000 | 0.677 | 0.583 | 0.787 |
| X-11444 levels | 0.002 | 1.117 | 1.041 | 1.198 |
| X-11470 levels | 0.025 | 1.084 | 1.010 | 1.163 |
| X-11880 levels | 0.017 | 0.836 | 0.721 | 0.969 |
| X-12117 levels | 0.029 | 1.091 | 1.009 | 1.180 |
| X-12261 levels | 0.046 | 0.863 | 0.746 | 0.997 |
| X-12822 levels | 0.015 | 0.883 | 0.799 | 0.976 |
| X-13553 levels | 0.042 | 0.877 | 0.772 | 0.995 |
| X-16397 levels | 0.018 | 1.280 | 1.044 | 1.571 |
| X-18901 levels | 0.038 | 0.844 | 0.720 | 0.990 |
| X-18913 levels | 0.015 | 1.209 | 1.037 | 1.409 |
| X-21807 levels | 0.008 | 0.863 | 0.774 | 0.962 |
| X-25371 levels | 0.001 | 1.194 | 1.077 | 1.324 |
| X-25422 levels | 0.008 | 0.866 | 0.778 | 0.964 |
| X-25828 levels | 0.003 | 1.420 | 1.123 | 1.794 |

**Table 2.** Enrichment analysis results of the causal relationship between plasma metabolites and fibromyalgia based on the KEGG, Reactome, Wikipathways and RaMP-DB database.

| Pathways | Total | Expected | P-value | FDR | Related metabolites |
| --- | --- | --- | --- | --- | --- |
| **KEGG** |  |  |  |  |  |
| Caffeine metabolism | 10 | 0.065 | 2.32E-05 | 0.00185 | 5-Acetylamino-6-formylamino-3-methyluracil, 7-Methylxanthine, 1-Methylxanthine |
| Ascorbate and aldarate metabolism | 9 | 0.0585 | 0.0571 | 1 | Glucuronate |
| alpha-Linolenic acid metabolism | 13 | 0.0845 | 0.0816 | 1 | EPA, Stearidonic acid |
| **Reactome** |  |  |  |  |  |
| Synthesis, secretion, and inactivation of Glucagon-like Peptide-1 | 21 | 0.08805 | 0.000062187 | 0.0011464 | D-Mannose, EPA, Oleoyl-EA |
| Incretin synthesis, secretion, and inactivation | 21 | 0.08805 | 0.000062187 | 0.0011464 | D-Mannose, EPA, Oleoyl-EA |
| Peptide hormone metabolism | 55 | 0.23061 | 0.0011464 | 0.41844 | D-Mannose, EPA, Oleoyl-EA |
| **Wikipathways** |  |  |  |  |  |
| Caffeine and theobromine metabolism | 11 | 0.065967 | 0.000026497 | 0.0074458 | 5-Acetylamino-6-formylamino-3-methyluracil, 7-Methylxanthine, 1-Methylxanthine |
| Biomarkers for urea cycle disorders | 22 | 0.13193 | 0.007127 | 1 | D-Mannose, Homocitrulline |
| Omega-3 / omega-6 fatty acid synthesis | 38 | 0.22789 | 0.020562 | 1 | EPA, Stearidonic acid |
| **RaMP-DB** |  |  |  |  |  |
| Caffeine and theobromine | 11 | 0.036 | 4.53E-06 | 0.015 | 5-Acetylamino-6-formylamino-3-methyluracil, |
| metabolism |  |  |  |  | 7-Methylxanthine, 1-Methylxanthine |
| Caffeine Metabolism | 19 | 0.0621 | 2.62E-05 | 0.0434 | 5-Acetylamino-6-formylamino-3-methyluracil,<br>7-Methylxanthine, 1-Methylxanthine |
| alpha-Linolenic acid metabolism | 18 | 0.0589 | 0.00147 | 1 | EPA, Stearidonic acid |
| Synthesis, secretion, and<br>inactivation of Glucagon-like<br>Peptide-1 | 21 | 0.0687 | 0.00201 | 1 | EPA, Oleoyl-EA |
| Incretin synthesis, secretion, and<br>inactivation | 21 | 0.0687 | 0.00201 | 1 | EPA, Oleoyl-EA |

KEGG: Kyoto Encyclopedia of Genes and Genomes; RaMP-DB: The Resource for Metabolite Pathway Database; EPA:Eicosapentaenoic acid; Oleoyl-EA:Oleoyl ethanolamide.

Using the identified metabolites, we conducted pathway enrichment analyses through the KEGG, Reactome, Wikipathways, and RaMP-DB databases. The results are shown in **Figure 4 and Table** 1. **2**. The primary metabolic pathways identified were Caffeine metabolism, Alpha-linolenic acid metabolism, Synthesis, secretion, and inactivation of Glucagon-like Peptide-1 (GLP-1), and Incretin synthesis, secretion, and inactivation (Screening criteria: [P_FDR_ < 0.05; [P < 0.05, P_FDR_ > 0.05, but the pathway is enriched in at least two out of the four databases: KEGG, Reactome, Wikipathways, and RaMP-DB. In the Caffeine metabolism pathway, FM-associated metabolites included 5-Acetylamino-6-formylamino-3-methyluracil (AFMU), 7-Methylxanthine, and 1-Methylxanthine. For the Alpha-linolenic acid metabolism pathway, Eicosapentaenoic acid (EPA) and Stearidonic acid (SDA) were identified. In the Synthesis, secretion, and inactivation of GLP-1 pathway, D-Mannose, EPA, and Oleoyl-EA were implicated, while the Incretin synthesis, secretion, and inactivation pathway similarly involved D-Mannose, EPA, and Oleoyl-EA.

**Figure 4.**
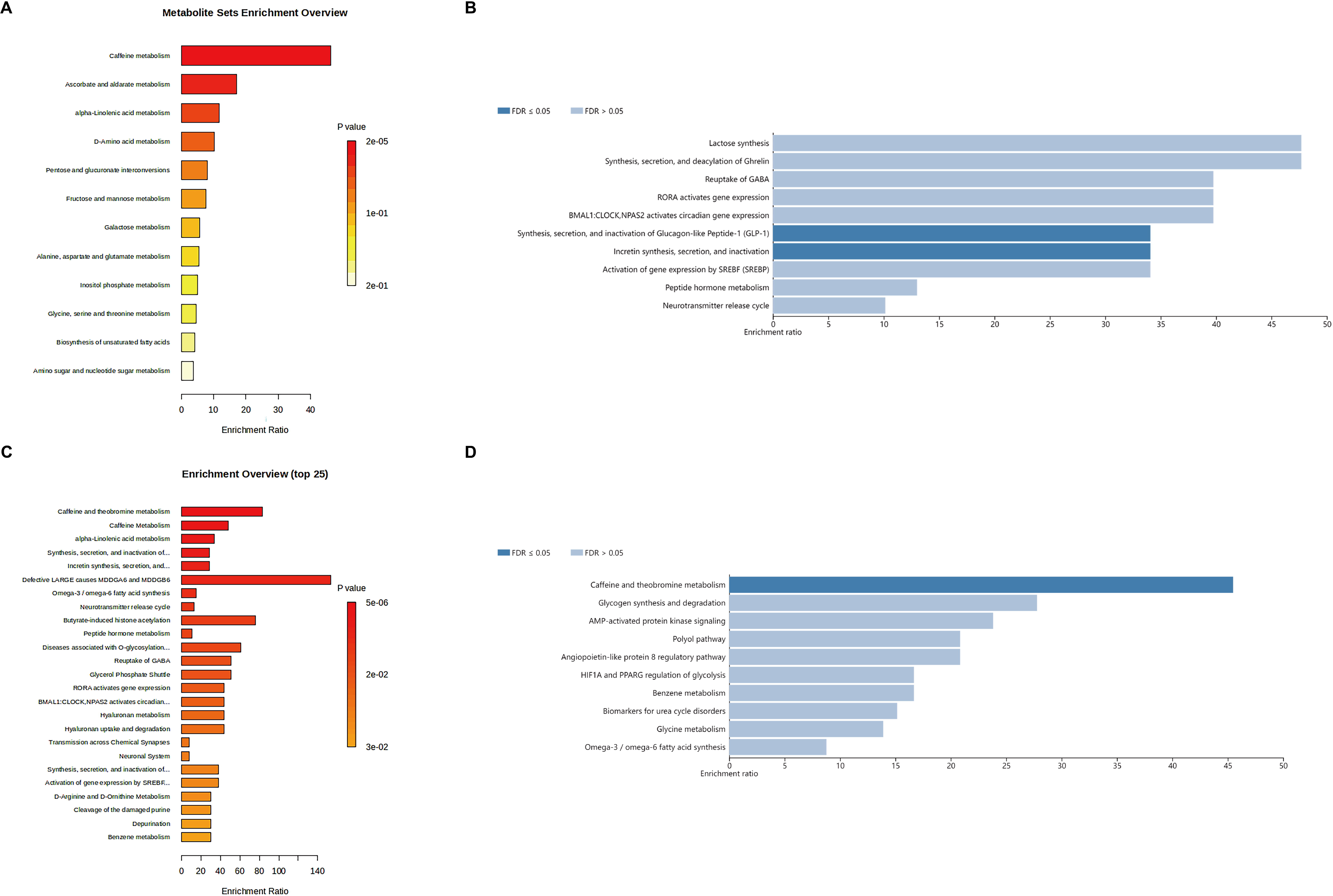
(A) Enrichment analysis results of the causal relationship between plasma metabolites and fibromyalgia based on the Kyoto Encyclopedia of Genes and Genomes (KEGG) database. (B) Enrichment analysis results of the causal relationship between plasma metabolites and fibromyalgia based on the reactome. (C) Enrichment analysis results of the causal relationship between plasma metabolites and fibromyalgia based on the RaMP-DB (Resource for Metabolite Pathway Database). (D) Enrichment analysis results of the causal relationship between plasma metabolites and fibromyalgia based on the wikipathways.

##### 3.3 MR Results for Gut Microbiota and Plasma Metabolites

We conducted MR analysis between six gut microbiota taxa and 82 plasma metabolites. Detailed results are available in **Table S2**. The analysis indicated causal relationships between family *Enterobacteriaceae* and several metabolites, including 2,3-dihydroxypyridine, 1-stearoyl-2-oleoyl-GPE (18:0/18:1), palmitoylcholine, and the threonine to alpha-ketobutyrate ratio. Additionally, genus *Butyricicoccus* was associated with sphingadienine, X-11444, X-16397, and the androsterone glucuronide to etiocholanolone glucuronide ratio. Genus *Coprococcus1* showed a correlation with (16 or 17)-methylstearate (a19:0 or i19:0), while genus *Eggerthella* was linked to glucuronate and 1-(1-enyl-stearoyl)-GPE (p-18:0) levels. Genus *Ruminococcaceae* UCG005 was associated with stearidonate (18:4n3), oleoyl ethanolamide, and the adenosine 5’-monophosphate (AMP) to glutamate ratio. Order *Enterobacteriales* was similarly associated with 2,3-dihydroxypyridine, 1-stearoyl-2-oleoyl-GPE (18:0/18:1), palmitoylcholine, and the threonine to alpha-ketobutyrate ratio.

##### 3.4 Mediation Effect

Based on the identified gut microbiota and plasma metabolites, multivariable MR analysis was performed to explore the mediating role of plasma metabolites in the impact of gut microbiota on FM, with results summarized in **Table 3**. Specifically, 2,3-dihydroxypyridine and palmitoylcholine levels were identified as potential mediators in the causal pathway between family *Enterobacteriaceae*, order *Enterobacteriales*, and FM, with mediation proportions of 14.91% and 12.34%, respectively.

**Table 3.** Mediation MR analysis results.

| **Exposure** | **Mediator** | **Outcome** | **Mediation Effect** | **Lower 95% CI** | **Upper 95% CI** | **Mediated proportion (%)** |
| --- | --- | --- | --- | --- | --- | --- |
| Family Enterobacteriaceae | 2,3-dihydroxypyridine | Fibromyalgia | -0.058 | -0.144 | -0.001 | 14.91% |
| Family Enterobacteriaceae | Palmitoylcholine | Fibromyalgia | -0.048 | -0.120 | 0.001 | 12.34% |
| Order Enterobacteriales | 2,3-dihydroxypyridine | Fibromyalgia | -0.058 | -0.144 | -0.001 | 14.91% |
| Order Enterobacteriales | Palmitoylcholine | Fibromyalgia | -0.048 | -0.120 | 0.001 | 12.34% |

##### 4. Discussion

This study utilized MR analysis to explore the causal relationships among gut microbiota, plasma metabolites, and FM. The findings underscore the significant roles of specific gut microbiota in FM pathophysiology, identifying four taxa—family *Enterobacteriaceae*, genus *Butyricicoccus*, genus *Coprococcus1*, and order *Enterobacteriales*—with protective effects, and two taxa—genus *Eggerthella* and genus *Ruminococcaceae*—as potential risk factors. Moreover, the study uncovered 82 plasma metabolites associated with FM, pointing to key metabolic pathways such as caffeine metabolism, α-linolenic acid (ALA) metabolism, and the synthesis, secretion, and inactivation of GLP-1 and incretin. Of particular note, *Enterobacteriaceae* and *Enterobacteriales* was shown to influence FM through the metabolites 2,3-dihydroxypyridine and palmitoylcholine, which act as mediators in the causal pathway. These results deepen our understanding of the interplay between gut microbiota and host metabolic pathways in FM and suggest promising directions for clinical interventions and future research.

#### 4.1 Gut Microbiota and Fibromyalgia

The association between gut microbiota and FM has been thoroughly explored in our previous studies. By integrating multi-omics data and advanced analytical methods, we have preliminarily elucidated the potential roles of specific gut microbiota in the pathophysiological mechanisms of FM ^28^.

#### 4.2 Plasma Metabolites and Fibromyalgia

This study identified 82 plasma metabolites associated with FM. Metabolic enrichment analysis revealed that these metabolites are primarily involved in caffeine metabolism, ALA metabolism, and the pathways responsible for the synthesis, secretion, and inactivation of GLP-1. Additionally, these metabolites are linked to the regulation of incretin metabolism. Disruptions in these metabolic pathways may constitute key features of the underlying pathophysiology of FM.

##### 4.2.1 Caffeine metabolism

This study found that the caffeine metabolism pathway may be involved in the pathogenesis of FM through specific metabolites, including AFMU (protective), 7-Methylxanthine (risk factor), and 1-Methylxanthine (risk factor). Caffeine exerts its effects by antagonizing adenosine receptors, which interferes with the key physiological functions of adenosine in pain regulation, anti-inflammation, and improving microcirculation and energy metabolism in muscles ^29–31^. 1-Methylxanthine and 7-Methylxanthine are methylxanthine intermediates of caffeine metabolism, produced during demethylation mediated by the key enzyme CYP1A2 ^32^. These compounds exhibit partial adenosine receptor antagonism, which may underlie their role as risk factors in FM ^33^. The metabolism of 7-Methylxanthine and 1-Methylxanthine is dependent on xanthine oxidase, a process associated with the generation of ROS and oxidative stress. The latter can impair mitochondrial function, reduce ATP production efficiency, and lead to muscle fatigue, disordered energy metabolism, and the chronic pain commonly experienced by FM patients^34–38^.

AFMU is one of the major end-products of caffeine metabolism and does not exhibit adenosine receptor antagonistic activity ^39^. Its formation involves multiple enzymes, including N-acetyltransferase 2 (NAT2). Polymorphisms in NAT2 are a major determinant of individual variability in caffeine metabolism ^40^. Elevated AFMU levels may indicate rapid clearance of caffeine metabolites, which helps prevent prolonged inhibition of adenosine signaling. Restoring adenosine’s analgesic and anti-inflammatory effects may alleviate symptoms in FM patients. Additionally, AFMU is produced via a non-oxidative pathway catalyzed by NAT2, which avoids ROS generation, thereby playing a protective role in FM.

In summary, abnormalities in caffeine metabolism, particularly those affecting adenosine signaling, may be a core mechanism underlying FM. The combined presence of low levels of the protective metabolite AFMU and high levels of the risk-associated metabolites 7-Methylxanthine and 1-Methylxanthine suggests a strong association between caffeine metabolism dysregulation and FM-related symptoms, including pain sensitization, neuroinflammation, and fatigue.

##### 4.2.2 **α**-linolenic acid metabolism

This study highlights the role of the ALA metabolic pathway, particularly through the protective metabolites EPA and SDA, in providing new insights into the pathogenesis of FM. ALA, an essential n-3 polyunsaturated fatty acid, is primarily found in plant oils such as flaxseed, walnut, and canola oil. Through enzymatic reactions, ALA is converted into SDA, EPA, and docosahexaenoic acid (DHA), which play significant roles in inflammation regulation, mitochondrial function improvement, and the restoration of neuro-immune balance. As explained in the Introduction, a lowered omega-3/omega-6 ratio is inversely associated with fibromyalgia symptoms in CFS ^15^. (1) Inflammation Regulation: EPA, DHA, and SDA are anti-inflammatory n-3 fatty acids that compete with the cyclooxygenase (COX) and lipoxygenase (LOX) pathways to reduce the production of pro-inflammatory mediators derived from ω-6 fatty acids, thus modulating inflammatory responses ^41,42^. EPA and DHA serve as precursors for specialized pro-resolving mediators, such as resolvins and protectins, which promote inflammation resolution and tissue homeostasis ^43,44^. Additionally, EPA and DHA incorporation into cell membranes alters membrane fluidity and protein functionality, influencing the production of arachidonic acid metabolites ^45,46^. These fatty acids also inhibit NF-κB activity and regulate the expression of inflammation-related genes, such as COX-2, thereby reducing the release of pro-inflammatory cytokines and mediators ^46–48^. Moreover, EPA and DHA alleviate oxidative stress by reducing ROS production and improving insulin resistance and metabolic health through the Mitogen-Activated Protein (MAP) kinase and GPR120/PPARγ pathways ^49,50^. (2) Mitochondrial Function Improvement: DHA supplementation enhances mitochondrial oxygen consumption in skeletal muscle and improves energy-sensing pathways, thereby boosting muscle function ^51^. EPA and DHA protect mitochondrial function by reducing oxidative stress and ameliorating mitochondrial dysfunction in metabolic disorders ^52^. Notably, EPA appears to have a greater advantage over DHA in restoring mitochondrial function, as it increases mitochondrial membrane potential and cardiolipin levels in astrocytes ^53^. (3) Neuro-Immune Balance Restoration: EPA and DHA improve neuroinflammation and neurodegenerative conditions by modulating neuroimmune pathways, altering membrane function, and competing with n-6 PUFAs ^54^. DHA, through interactions with the GPR120 receptor, regulates immune cell function and modulates the Th17/Treg balance during viral infections ^55^.

Additionally, supplementation with n-3 PUFAs has shown positive effects in improving depression and metabolic abnormalities^56^. For example, in a study involving 24 patients with major depressive disorder (MDD), EPA treatment significantly increased activity in brain regions related to emotion perception and cognitive control, suggesting a potential advantage of EPA in alleviating depressive symptoms ^57^. Moreover, diets rich in EPA phospholipids have been shown to mitigate chronic stress and LPS-induced depression- and anxiety-like behaviors by regulating the immune system and neuroinflammation ^58^. In patients with CFS, EPA and DHA supplementation has demonstrated potential for improving metabolic dysfunction. Studies have shown that muscle cells in CFS patients fail to effectively activate the Adenosine Monophosphate-Activated Protein Kinase (AMPK) pathway or increase glucose uptake after exercise. Supplementation with EPA and DHA helps address these metabolic deficits and restores energy metabolism balance ^59^.

In conclusion, the pathogenesis of FM may involve low-grade chronic inflammation, oxidative stress, energy metabolism abnormalities, and impaired muscle function. Regulation of the ALA metabolic pathway offers new perspectives for understanding FM mechanisms and therapeutic interventions.

##### 4.2.3 Synthesis, secretion, and inactivation of glucagon-like peptide-1 and incretin

This study identified EPA (a protective omega-3 PUFA) and Oleoyl-EA (a risk-related metabolite) as key regulators in the GLP-1 and incretin synthesis, secretion, and inactivation pathways, providing novel insights into the pathogenesis of FM. Previous research has demonstrated that GLP-1 improves insulin sensitivity and regulates blood glucose levels, helping patients achieve metabolic homeostasis and alleviating metabolic disorders ^60^. Additionally, GLP-1 enhances glucose uptake in muscle tissues and optimizes ATP production. It also activates antioxidant enzymes, reducing free radical levels and mitigating oxidative stress damage ^61^. This is essential for protecting mitochondria from oxidative damage, maintaining muscle function, and preserving energy metabolism balance. Furthermore, GLP-1 suppresses the release of pro-inflammatory cytokines, modulates immune responses, and reduces the impact of chronic inflammation on the body ^62^. These functions position GLP-1 as a critical target for FM.

EPA, as an omega-3 polyunsaturated fatty acid, promotes GLP-1 synthesis and secretion by regulating cell membrane fluidity and signaling pathways ^63^. It may also inhibit dipeptidyl peptidase-4 (DPP-4), the primary enzyme responsible for GLP-1 degradation, thereby prolonging the biological activity of GLP-1 and enhancing its anti-inflammatory and antioxidant effects ^64,65^. In contrast, Oleoyl-EA, a risk-associated metabolite, may reduce GLP-1’s protective effects by promoting DPP-4 activity or interfering with GLP-1 secretion. The precise mechanisms underlying Oleoyl-EA’s actions require further investigation.

In summary, the study of the GLP-1 pathway and its metabolites may further elucidate FM’s pathogenesis. The protective effects of EPA and the detrimental impact of Oleoyl-EA suggest that metabolic pathway dysregulation and its interplay with inflammation and oxidative stress may represent critical mechanisms underlying FM.

##### 4.2.4 Oxalate and FM

This study suggests that oxalate may be a potential risk factor for FM, although no direct evidence currently establishes a definitive link. Previous research indicates that FM patients often experience other metabolic disorders, such as hyperuricemia, which may be associated with low-grade inflammation induced by urate crystals ^66^. Furthermore, chronic inflammation, neuroendocrine dysfunction, and other metabolic factors are known to play critical roles in the pathogenesis of FM ^67^. These findings suggest that FM involves multiple pathophysiological mechanisms, and the role of oxalate as a metabolic byproduct warrants further investigation and attention.

##### 4.2.5 Dietary Recommendations for FM

This study highlights the importance of dietary adjustments for FM patients to regulate metabolic pathways, reduce inflammation and oxidative stress, and alleviate symptoms. The following dietary strategies are proposed: (1) Caffeine Management: Patients should limit caffeine intake to prevent pro-inflammatory effects from metabolic intermediates such as 7-Methylxanthine and 1-Methylxanthine. (2) Omega-3 Fatty Acids: Increase the intake of omega-3 fatty acid-rich foods, such as flaxseed oil, walnuts, oily fish, and fish oil, to promote the production of protective metabolites like EPA and SDA. Concurrently, reduce omega-6 fatty acid consumption to balance the fatty acid ratio. (3) Low Glycemic Index (GI) Diet: Adopt a low-GI diet by choosing whole grains and high-fiber foods to promote GLP-1 secretion. Limit the consumption of high-fat foods rich in Oleoyl-EA to mitigate its adverse effects on inflammation and metabolism. (4) Oxalate Reduction: Reduce intake of high-oxalate foods, such as spinach and beets, and consider moderate calcium supplementation to lower the metabolic burden of oxalate. Under medical supervision, consider supplementing with omega-3 fatty acids and antioxidants to combat low-grade chronic inflammation, metabolic dysregulation, and neuroinflammation in FM. In conclusion, personalized dietary interventions, including nutrient optimization and anti-inflammatory strategies, can help mitigate symptoms associated with FM, such as chronic low-grade inflammation, energy metabolism disorders, and neuroinflammation.

#### 4.3 Gut Microbiota-Metabolites-FM

This study identified that *Enterobacteriaceae* and *Enterobacteriales* influences FM through plasma metabolites, with 2,3-dihydroxypyridine and palmitoylcholine serving as mediators in the causal pathway. Recent research has highlighted the significant role of gut microbiota and their metabolites in the pathogenesis of FM. Specifically, members of the gut microbial family *Enterobacteriaceae* affect host physiological functions via their metabolic products ^68^. Studies suggest that 2,3-dihydroxypyridine, a key intermediate in iron metabolism, is closely associated with oxidative stress responses in the host ^69,70^. Similarly, palmitoylcholine, a derivative of the choline metabolism pathway, plays a role not only in fatty acid synthesis but also in regulating metabolic networks through signaling mechanisms ^71,72^. These two metabolites mediate the interaction between gut microbiota and the host, influencing inflammation, oxidative stress, and neurotransmitter metabolic pathways ^73,74^. Consequently, they may impact pain and fatigue symptoms in FM patients, providing novel insights into the mechanisms underlying FM development.

### 4.4 Limitations

This study is the first to apply a comprehensive MR framework to analyze the causal relationships between gut microbiota, plasma metabolites, and FM. Furthermore, multivariable MR and mediation analyses through plasma metabolites were employed to construct the pathway linking gut microbiota to FM. Sensitivity analyses were conducted to maximize the robustness of MR results. However, this study has certain limitations. First, the lack of demographic information, such as age and gender, in the initial datasets prevented further subgroup analyses. Second, the majority of the individuals included in the GWAS datasets were of European ancestry, which limits the generalizability of the findings to other populations. Lastly, while the MR method is effective for evaluating causal relationships between exposures and outcomes, these results need further validation through additional experimental and clinical studies.

### 5. Conclusion

In conclusion, this study systematically assessed the causal relationships between gut microbiota, plasma metabolites, and FM using MR analysis. The results highlight the protective roles of four gut microbial taxa (family *Enterobacteriaceae*, genus *Butyricicoccus*, genus *Coprococcus1*, and order *Enterobacteriales*) and identify two taxa (genus *Eggerthella* and genus *Ruminococcaceae*) as potential risk factors. Additionally, 82 plasma metabolites associated with FM were identified, with significant involvement in pathways such as caffeine metabolism, ALA metabolism, and GLP-1 and incretin regulation. The mediating effects of 2,3-dihydroxypyridine and palmitoylcholine further elucidate how *Enterobacteriaceae* and *Enterobacteriales* contribute to FM pathogenesis. Moreover, personalized dietary interventions—including limiting caffeine intake, increasing omega-3 fatty acid consumption, adopting a low GI diet, and reducing high-oxalate foods—may help regulate metabolic pathways, reduce inflammation and oxidative stress, and alleviate FM-related symptoms. This study not only underscores the intricate interactions between gut microbiota and metabolic pathways but also proposes actionable targets for clinical intervention, dietary management, and precision medicine approaches in FM.

### Ethics approval and consent to participate

Our analyses were based on publicly available data that had been approved by relevant review boards. Therefore, no additional ethical approval is required.

### Consent for publication

N/A.

### Funding

There was no specific funding for this specific study.

## Data Availability

The datasets used and/or analyzed during the current study are presented in the manuscript. Summary statistics for GWAS are publicly available.

## Acknowledgment

The research community is thanked for making summary statistics from genome-wide association studies publicly available.

## Author’s contributions

Mengqi Niu: conceptualization; data curation; formal analysis; writing - original draft. Jing Li: data curation, writing - review & editing. Youman Zhuang: data curation. Chenkai Yangyang: data curation. Yali Chen: data curation. Yingqian Zhang: supervision; writing - review & editing. Michael Maes: conceptualization; supervision; writing - review & editing. All authors approved the submitted manuscript.

